# A Systematic Review of Evidence Behind the CDC Guidelines for Indoor Lightning Safety

**DOI:** 10.1101/2023.10.05.23296621

**Authors:** Elissa Brown, Alyson Haslam, Vinay Prasad

## Abstract

**Objective:** To assess the evidence underlying the Centers for Disease Control and Prevention (CDC) indoor safety guidance for lightning storms.

**Design:** Systematic Review of peer-reviewed literature.

**Setting:** Google Scholar, PubMed, and Web of Science (through 2023).

**Participants:** Reports of indoor death or injury from lightning strike.

**Main outcome measures:** The number of deaths and injuries from lightning-related activities.

**Results:** A majority of the 15 articles identified were retrospective reviews of data from death certificates, medical records, and newspaper reports; 5 articles were based on single case reports. Reports of injuries from lightning while indoors are exceedingly rare; death from lightning while indoors is essentially non-existent in modern times. No evidence exists that supports the given advice.

**Conclusions:** Current U.S. lightning avoidance tips may inadvertently portray indoor spaces as unsafe, despite their protective advantage. Guidelines should place less emphasis on indoor situations and highlight controllable risks, such as behavior in outdoor and recreation situations.

## Introduction

The Centers for Disease Control and Prevention (CDC) in the United States (U.S.) has issued guidance on indoor lightning injury, which has most recently been updated in 2022. They caution that a third of all lightning injuries occur indoors.^1^ To minimize this risk, the CDC advises several precautions. First, avoid water-related activities like bathing and dishwashing, as lightning can exploit plumbing pathways. Second, refrain from using electronic devices linked to outlets, since lightning can traverse electrical systems and metal elements. Third, steer clear of windows, doors, and concrete surfaces to avoid metal-conductive pathways. Finally, avoid corded phones during thunderstorms, due to potential conductivity.^1^ These recommendations can be juxtaposed against the absolute risk of lightning injury among Americans.

According to CDC advice, Americans should not do dishes or shower during thunderstorms, and should refrain from standing near any windows. While less than 2% of U.S. households rely solely on a landline phone (cite), the CDC recommends avoiding corded phones. Given this advice, we sought to assess the absolute risk of injury while engaging in these activities to determine whether the CDC indoor lightning advice is sensible or whether it should be focused on tips to avoid activities, such as outdoor recreation, that would ultimately have greater impact.

Lightning strikes the earth approximately 8 million times a day or 100 times per second.^2^ According to the National Oceanic and Atmospheric Administration, lightning kills an average of twenty people per year in the U.S..^3^ Climate, region, and season play a role in the risk of lightning injury, as shown by the differences in incidence of lightning injury across the U.S.; Florida and Texas account for the most lightning fatalities, while Alaska, Delaware, Hawaii, New Hampshire, Washington and the U.S. Virgin Islands have reported no lightning deaths since 2006.^4^

Despite considerable population growth, there are far fewer annual lightning fatalities in the U.S. compared to the previous century.^5^ In the 1920s–1940s, the U.S. experienced between 300 and 400 lightning deaths per year while the average from 2013 to 2022 was just 22 deaths per year.^5^ On an individual basis, this decline in lightning deaths in the U.S. is staggering, falling from about 3 or 4 annual deaths per million, to fewer than 0.1 death per million in the past 10 years.^6^ Experts generally agree that a combination of urbanization and modernization have played a significant role in decreasing death from lightning.

This review aims to determine the empirical data underlying the CDC’s indoor lightning safety guidelines by assessing the available evidence on death and injury due to lightning strikes while indoors.

## Methods

We sought to assess the evidence for lightning storm safety guidance by compiling a list of articles reporting on death and injury during indoor activities related to lightning strikes. In compiling a list of relevant activities to include, we reviewed the CDC’s lightning safety tips for indoor activities on which the CDC has made specific guidance.^1^

### Search strategy

We searched PubMed and Web of Science using iterations of “phones, bathing, computers, or electrical cords” and “lightning”. In addition, we searched Google Scholar, using the phrase “activities of death or injury during lightning storm” and retrieved articles using Publish or Perish software.^10^ We used the first 100 search results. The search date was August 8, 2023.

We included articles that were primary reports of death or injury from indoor activities related to lighting strikes. If an article was published more than once, we reviewed the first publication if accessible, otherwise we included the version with full-text availability. We excluded articles reporting only lightning statistics for outdoor activities, where indoor vs. outdoor location was not specified, articles without information on activities or location relating to the strike, articles reporting on physiology or medical outcomes of lightning effects, mechanics of lightning strikes, climatology, or attitudes/beliefs.

We also excluded articles discussing sports-related activities as these are not covered by the CDC’s guidance. We reviewed commentary and articles for other relevant citations that discuss death or injury from activities related to lightning strikes.

### Data abstraction

For our analysis, we abstracted relevant activities in accordance with the four CDC prevention strategies as those relating to talking on corded phones, activities requiring the use of equipment with electrical cords (e.g., computers), activities involving plumbing (e.g., washing dishes, showering/bathing), and activities involving being near windows, doors, or concrete surfaces. We also abstracted data on the country or geographical region, years of data ascertainment, data source, study type, number of incidents (deaths and/or injuries), age of participants, and death rate (if reported). We omitted any data originating before the 20^th^ century. Data were abstracted by two authors (EB and AH).

To get a sense of death and injury rate if not reported in the article we searched the United Nations – World Population Prospects for population counts for the years of study observation.^11^ Since most death and injury counts were an average of the study observation, we used the median population for the study years and divided the study mortality rate by the number of years to get the yearly mortality rate. Rates were calculated per one million people.

We included individual case reports in our pooled estimates and conducted a narrative review for select individual case studies to highlight unusual settings and outcomes.

### Statistical analysis

Data were presented as descriptive frequencies. All data were analyzed with R statistical software version 4.1.1 (R Foundation).

In accordance with 45 CFR §46.102(f), Our study was not submitted for institutional review board approval because it included publicly available data and did not involve individual patient data.

## Results

Among search results, we found 12 papers that met our inclusion criteria. Our review uncovered an additional 3 articles (all case reports) that met our inclusion criteria, for a total of 15 relevant articles reporting on activities related to deaths and injuries from lightning while indoors.

The majority of articles (n=9; 60.0%) were published in the 21^st^ century; only one was published before 1990. The years of study span from 1891 through 2016 and reflect a total of 7,296 incidents. Incidents from prior to 1900 were omitted from this analysis (n= 1,043) for a total analytic sample of 6,253 incidents of lightning injury or death, with a median of 121 incidents (IQR: 352.75); 20.0% of all incidents occurred indoors (n=1,251) and 80% (n=5,002) occurred outdoors or the location was not known (**Figure**). There is notable overlap in the data from the U.S.

**Figure.**
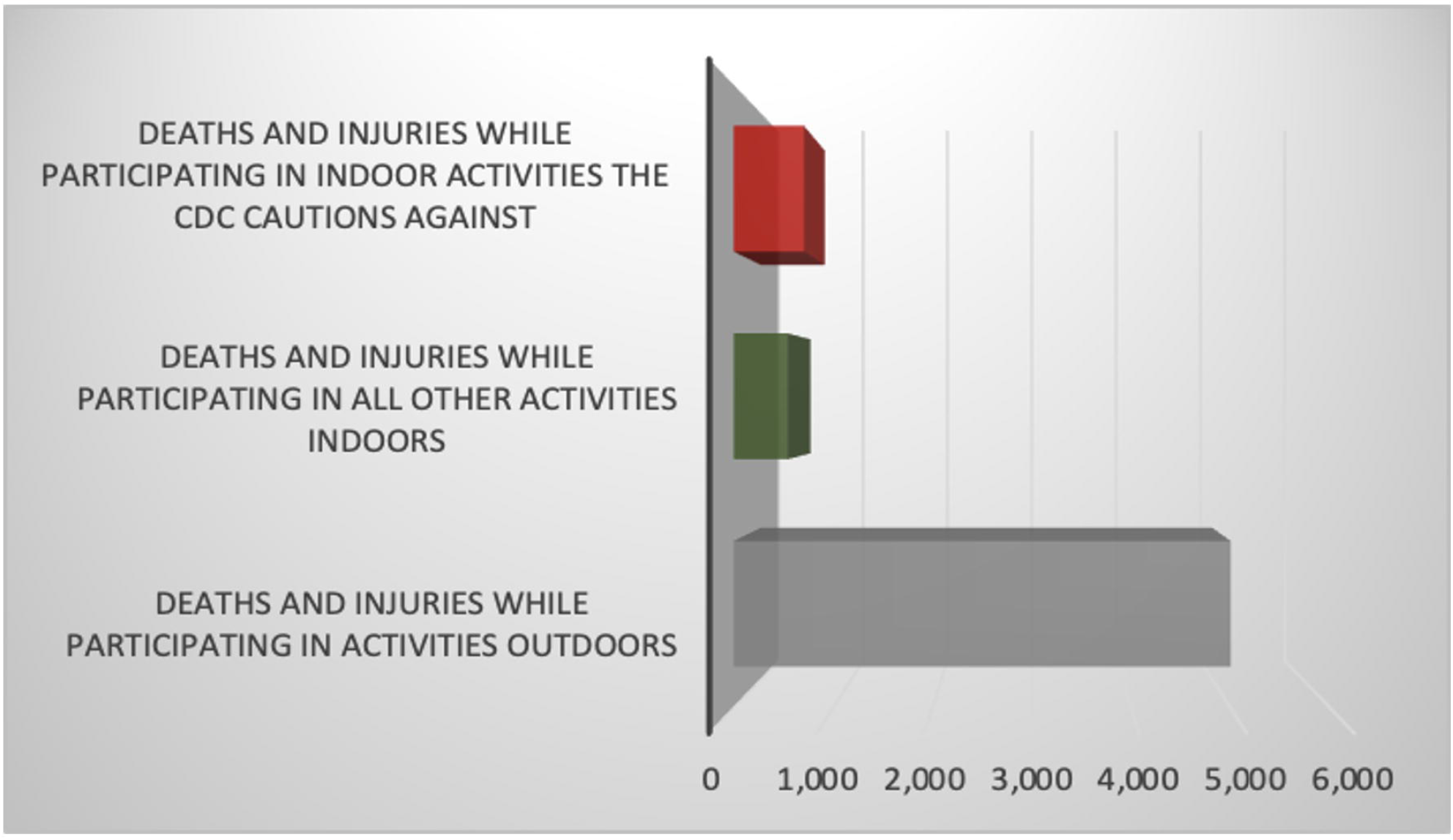
Incidents of lightning death and injury during activities cautioned against by the CDC, compared to other indoor activities and outdoor activities.

Five articles (33.3%) represent individual case reports and 10 articles (66.7%) are retrospective reviews of death certificates, medical records, and newspaper reports. The median number of injuries and deaths are 0 (IQR: 246) and 31 (IQR: 76.5), respectively. Reports on lightning injuries and deaths in the U.S. made up the largest percentage of articles (n=7; 46.7%), followed by the United Kingdom (U.K.; n=3; 20.0%). Reports on deaths and injuries from South Africa (Swaziland), Albania, and Australia each contributed one article (n=1; 6.7%), while 13.3% of articles did not specify a study location (n=2).

The median number of years included in each study was 7. The median annual death rate (per million) attributed to lightning strikes for indoor-related activities was 0.09 and ranged from 0 for Australia to 23 for South Africa (Swaziland).

### Corded phones

Our analytic sample includes 7 reports of death when struck by lightning while using a corded telephone, representing 0.4% of all deaths in our sample. All seven were reported in the U.S. Four of these incidents (57.1%) occurred between 1959 and 1965; two occurred between 1990 and 2010 and due to overlapping data it is uncertain whether these represent the same occurrence. Our sample does not include any reports of death by lightning while using a corded telephone since 2010.

There are 546 reports of injury from being struck by lightning while using a corded telephone in our dataset, representing 12.9% of all injuries in our sample. The majority of these injuries (n=373, 68.3%) occurred in Australia between 1980-1985; 18.9% occurred in the U.S. (n=103) from 1959-2010, and 12.8% occurred in the U.K. (n=70) from 1988-2012.

### Water-related activities

Our dataset includes 2 reports of death when struck by lightning while doing a water-related activity while indoors (e.g., taking a shower or washing dishes), representing 0.1% of all deaths in our sample. One was reported North Carolina (U.S.) between 1978 and 1988 with the decedent recorded as being “at sink”. The other was reported in Swaziland (South Africa) between 2003-2007 and recorded as “bathing” but it is unclear whether the decedent was indoors or outdoors. This incident should be interpreted in the context of the study setting; Swaziland is a country where a quarter of the population reside in stick and mud houses without plumbing.^12^ The study authors note that most fatalities occurred either outdoors or in unprotected, unsafe rural dwellings, the latter accounting for most fatalities (Dlamini 2009). Our sample does not include any reports of death while conducting an indoor water-related activity in the U.S. since 1988.

There are 30 reports of injury from lightning strikes while doing a water-related activity indoors in our dataset, representing 0.7% of all injuries in our sample. Half of the incidents (n=15) occurred in the U.K. between 1988-2012; fourteen incidents (46.7%) occurred in the U.S. between 1990-2010 and one incident occurred in the U.S in 2013.

### Electronic devices with cords

Our sample includes 10 reports of death when struck by lightning while using an electronic device with a cord while indoors, representing 0.6% of all deaths in our sample. All ten incidents occurred in the U.S. from 1978-1994. Our sample does not include any reports of death while using a corded device since 1994.

There are 80 reports of injury from lightning strikes while using a corded device indoors in our dataset, representing 1.9% of all injuries in our sample. The majority of these incidents (n=62, 77.5%) occurred in the U.S. between 1990-2010; due to overlapping data it is uncertain whether some of these represent the same occurrences. Regardless, our sample does not include any reports of injury while using a corded device indoors in the U.S. since 2010. The remainder of these incidents (n=18, 22.5%) occurred in the U.K. between 1988 and 2012.

### Windows, doors, and cement surfaces

Our dataset includes one report of death from being struck by lightning while near a door, window, or concrete surface while indoors, representing 0.1% of all deaths in our sample. This death occurred in the U.S. between 1990-2010. Our sample does not include any reports of death from being struck by lightning while standing near a door, window, or concrete surface indoors since 2010.

There are 30 reports of injury from lightning strikes while standing near a door, window, or concrete surface in our dataset, representing 0.7% of all injuries in our sample. The majority of these incidents (n=29, 96.7%) occurred in the U.S. between 1990-2010; due to overlapping data it is uncertain whether some of these represent the same occurrences. Regardless, our sample does not include any reports of injury while using a corded device indoors in the U.S. since 2010. The remaining incident occurred in the U.K. in 1992.

### Defining an indoor setting

Two case reports of death by lightning strike indoors should be reviewed in the context of the settings. Kreci et al describes the death of a 46-year-old Albanian male, caused by lightning striking the chimney of his home while he was touching the wood stove. The authors explain that the incident took place in a rural area where many Albanian families primarily heat their homes with wood stoves and have chimneys that are hand-made and non-professionally mounted – in this case the chimney is essentially a metal tube directly connecting the furnace with the outside.^13^ Ventura et al, does not provide a location but notes that the victim was doing renovation work on a cottage, which is described as a “rudimentary system of walls with steel beams connecting the exterior of the structure to the central portion of the cottage through the roof”.^14^

Finally, it should be noted that within our analytic sample we identified 3 incidents of injury and 3 incidents of death by lightning strike while in bed, representing 0.1% of all injuries and 0.2% of all deaths in our sample.

## Discussion

Over the past century, urbanization and significant advancements in infrastructure have made being indoors during a lightning storm remarkably safe. While there have been documented cases of people sustaining injuries due to lightning strikes even within well-grounded buildings over the last 30 years, the implementation of modern safety practices has resulted in a notable absence of fatalities. This is in stark contrast to a hundred years ago when the possibility of being struck by lightning while indoors was considerably more perilous. Contemporary measures like surge protectors, designed to reduce the potential electrical current exposure for individuals using electronic devices and computers, have contributed significantly to enhancing indoor lightning safety.^15^

Reported incidents of lightning-related injuries sustained indoors tend to be of a minor nature. For example, one individual case report occurred in July 2013 when a 47-year-old man was inside his home in Maui and unexpectedly witnessed a blue streak connecting the faucet to his hand while washing dishes.

Although the shock momentarily brought him to his knees, he managed to recount the incident to his wife in the living room shortly thereafter, reporting only five minutes of hand numbness.^16^ Furthermore, between 1993 and 1999, out of the 341 reported cases of individuals struck by lightning indoors in the U.K., the majority perceived the effects as relatively inconsequential and opted not to seek further medical attention.^17^

Despite the relative paucity of lightning related death for indoor activities and the minor nature of the injuries, the CDC issues guidance on how to further reduce this risk when inside during a thunderstorm. This guidance advises against showers and dishwashing, leading to potentially unhygienic and unsanitary conditions around the house, in an effort to avoid lightning death. We find little support that the absolute risk indoors is high, and no evidence that the advice is helpful. The work hours used to create the CDC’s guidance may far outweigh the life years gained from the advice.

The 20th century saw a significant shift in the U.S. population from rural to urban areas.^7^ Urbanization in the U.S. was accompanied by increased investment in infrastructure, including electrical grids and plumbing systems, which assist in dissipating lightning’s energy safely into the ground. These conductive pathways in buildings also reduce the risk of damage and fires, making homes and buildings less susceptible to the destructive effects of lightning strikes. Urban areas were often the first to benefit from electrical and plumbing advancements, but in 1935, the Rural Electrification Administration (REA) was established as part of President Franklin D. Roosevelt’s New Deal.^9^ With these improvements in infrastructure, the risk of injury or death from being indoors during a lightning storm has dropped to nearly zero, making historical guidance on lightning safety outdated and irrelevant.

These findings are limited in that reports on fatalities and injuries were not always clearly described, and we relied on the information that the authors presented in the articles. Also, while we searched several databases, we likely missed some reports, as many reports are not found in the peer-reviewed literature. As such, these results may be undercounts and not generalizable to the entire population. However, given that several articles were the most comprehensive collections of lightning reports in the media or hospital records, the incidents are likely to be the ones most relevant to informing public health policy.

## Conclusion

The National Lightning Safety Council says if you go inside before lightning becomes a threat, your personal odds of being struck are near zero.^18^ Yet, the CDC cautions that one-third of lightning injuries occur indoors and emphasizes the dangers of engaging in specific indoor activities.^1^ Considering the evidence, the CDC’s approach may inadvertently convey a message that being indoors is inherently unsafe and contradict the actual protective advantage offered by indoor spaces. A more effective approach to harm prevention would be to direct attention towards activities that harbor reasonable risks and are within people’s control.

Finally, recommending extreme measures to reduce a trivial level of risk, like not doing dishes during a thunderstorm to avoid being struck by lightning indoors, may undermine the CDC’s credibility on other issues. The public may also stop listening if they find the advice to be extreme or inconvenient without perceived benefit. While isolated incidents of lightning-related fatalities during indoor activities have been documented, it is vital to remember that the risk pales in comparison to many everyday activities we engage in without a second thought—activities that carry inherent risk even in the absence of lightning storms. It is essential to maintain a balanced perspective, avoiding the creation of an environment where individuals are immobilized by fear in their day-to-day actions. By understanding and contextualizing risks, the CDC can empower informed decision making without undue apprehension. This approach aligns with the core principles of risk management and responsible communication, encouraging the public to enjoy life while taking prudent precautions.

## Data Availability

All data produced in the present study are available upon reasonable request to the authors.

